# Financial Cost Analysis Associated with a Locomotor Exercise Randomized Controlled Trial in Chronic Stroke

**DOI:** 10.1101/2023.12.20.23300342

**Authors:** Emily M. Hazen, Bria L. Bartsch, Sandra A. Billinger

## Abstract

**Background:** Post-stroke recovery trials pose distinct recruitment and retention challenges, and understanding the financial requirements of conducting randomized controlled trials is crucial to ensure sufficient resources for successful study execution. The purpose of this analysis was to quantify the costs at a single site with a large catchment area of the Moderate-Intensity Exercise Versus High-Intensity Interval Training to Recover Walking Post-Stroke, HIT Stroke Trial.

**Methods:** To determine cost, study expense reports were gathered and divided into four categories: oversight, recruitment, retention, and outcome assessments. Categories were then further divided into chronological order for initial contact and prescreening, consenting, initial screening, and baseline testing. The 12-week intervention was divided into 4-week blocks: intervention block 1, post 4-week outcome testing, intervention block 2, post 8-week outcome testing, intervention block 3, and post 12-week outcome testing.

**Results:** Total direct cost for site execution was $539,768 with cost per participant approximated as $35,984. Oversight costs accounted for 65.8% of the budget at $355,661. To achieve goals related to inclusive participant recruitment ($21,923) and retention ($28,009), our site costs totaled $49,932. Direct study-related costs included screening assessments ($5,905), baseline assessments ($15,028), intervention ($76,952), and outcome assessments ($36,288).

**Conclusion:** Clinical trials on walking rehabilitation and exercise, especially those involving multiple assessment visits, require intensive oversight. This cost analysis provides important and critical insight into the expenses required to successfully execute an exercise-based walking rehabilitation trial in the United States.

## INTRODUCTION

A key issue surrounding stroke recovery trials is recruitment and retention of participants. Our previous work^1^ and others ^2–5^ have highlighted the trial barriers, demands on the study team, and proposed strategies for successful recruitment and retention. However, to our knowledge, exploring the cost associated with stroke recovery trials is non-existent. Recognizing the financial requirements of clinical trials is essential for ensuring success and facilitates effective planning and trial execution.^6–8^ Further, insight into cost demand allows for adequate funds to be devoted to resource procurement, study recruitment, outcome assessments, salaries, and other indirect study-related costs. The underestimation of required funds can hinder study execution and data quality, lead to wasted resources, and negatively impact the clinical decision-making which ultimately impacts patient care.^9,10^

Pharmaceutical trial costs have been well-documented, with a recent study reporting the average cost of a clinical drug trial was $48 million.^11^ Given the potential for robust impact on health and aging, exercise emerges as a powerful non-pharmacological alternative. Recognizing its status as “one of the most promising interventions to directly delay physiological decline and increase the health span,”^12^ understanding optimal exercise prescription parameters and their associated clinical trial costs becomes critical. A few studies have explored the cost demands for exercise trials in older adults.^13–15^ In particular, a phase III, multisite randomized controlled trial enrolling a targeted sample of 639 older adult participants reported the total cost to randomize one participant through study completion cost approximately $16,494 for an estimated total cost of $10.5 million dollars.^13^ By allocating sufficient resources to stroke rehabilitation and recovery interventions, particularly in the context of support from initiatives like the NIH StrokeNet^2^ and other funding mechanisms, we enhance scientific rigor and reproducibility in clinical trials with the aim of influencing clinical practice and public health outcomes including overall mortality.^16–19^ Documenting the cost demand of a walking rehabilitation exercise trial in chronic stroke will: 1) Provide increased insight into the financial resources required to successfully execute an exercise study, and 2) Allow future investigators to devote adequate finances to budget applications, study start-up, and study execution.

HIT-Stroke was a 12-week, National Institute of Health funded, multi-site exercise trial to determine the optimal training intensity for improving walking capacity in individuals 6-months to 5-years post-stroke. The detailed study protocol and main trial results have been published elsewhere.^16,20^ The purpose of this cost analysis was to report the cost demands associated with the University of Kansas Medical Center site. As we’ve published previously, our site serves a large catchment area that spans our suburban and rural areas of Kansas.^1^ Kansas City is automobile-centric with poor availability of public transportation,^21^ which limits opportunities for research participation at an academic medical center. Our study team strives for inclusive science practices in our clinical trials and a cost analysis would provide insight into the reality of successful trial execution, including transportation costs. Here, we provide a detailed summary of costs associated with: 1) Resource procurement, 2) Recruitment, 3) Intervention delivery, 4) Outcome assessments, 5) Salaries, and 6) Indirect study-related costs, such as participant transportation. Further, we provide insight into budgeting to overcome common stroke recovery and rehabilitation research barriers, such as affordable transportation, parking considerations, navigating from parking to laboratory, and treatment compliance.^2^

## METHODS

Licensed physical therapists completed intervention and assessment training and completed the required competencies for their respective roles (intervention or blinded assessor). Outcome assessments occurred across the 12-week intervention at baseline, 4, 8, and 12 weeks. Prior to randomization, participants successfully completed: (1) written informed consent, a medical history and medical record review, (2) the Patient Health Questionnaire (PHQ-9), (3) a 2-step command, (4) lower extremity Fugl-Meyer Motor Assessment, (5) lower limb spasticity assessment (Ashworth Scale), and (6) NIH Stroke Scale ataxia and neglect items.

At baseline, participants completed blinded assessments including: (1) a pre-visit form with repeated blood pressure measurements, (2) 10 meter walk tests at both comfortable and fastest possible speeds,^22^ (3) a 6-minute walk test,^23^ (4) functional ambulation category, (5) the EuroQol-5D (EQ-5D), (6) Activities-specific Balance Confidence (ABC) Scale, (7) Patient Reported Outcomes Measurement Information System (PROMIS) Fatigue Scale,^24^ (8) and a graded exercise test (GXT).^25^ These outcome assessments were repeated at 4, 8, and 12 weeks in addition to the Participant Ratings of Change survey. During the 12-week intervention, participants completed three 45-minute treatment sessions a week consisting of a 3-minute warm-up of overground walking, 10 minutes of overground training of either high-intensity interval training (HIT) or moderate aerobic training (MAT), 20 minutes of harness-assisted treadmill training (HIT or MAT), a second bout of 10-minute overground training (HIT or MAT), and a 2-minute cool down of overground walking. Lactate was collected once a week.

To estimate the financial costs of the HIT Stroke study at the University of Kansas Medical Center site, expenses were assigned to seven different categories: Recruitment, Screening Assessments, Baseline Assessments, Intervention, Outcome Assessments, Retention, and Oversight. Each of these categories includes costs related to study team effort, equipment and materials, and facilities and services used. Recruitment costs include factors such as staff time spent making recruitment calls and scheduling, newspaper advertisements, social media, and time spent introducing the study in clinic and stroke support groups. Screening Assessments include costs for training physical therapist time and assistance from students. Baseline Assessments include testing physical therapist time and space usage for graded exercise testing in addition to walking tests. Intervention consists of cost of equipment including initial purchases and maintenance, physical therapist time for intervention delivery, and student workers. Outcome assessments include costs of administration of graded exercise tests, testing physical therapist time, and required equipment for each assessment. Retention costs include participant compensation for successful completion of each outcome testing visit, transportation costs to and from study visits, and medical translator fees. Oversight includes principal investigator, study coordinator, and physical therapist efforts for the intervention delivery and the assessor (physical therapist) who was blinded to group assignment across trial duration. Oversight activities include initial study start up activities such as initial training, training site study staff, Data Safety and Monitoring Board meetings, adverse event reporting, data queries, maintaining supplies, overall study coordination, and regulatory document submissions to the Institutional Review Board (IRB).

To understand the distribution of these costs across the timeline of the trial, expense categories were further subdivided into chronological groups: Initial Contact and Prescreening, Consenting and Screening, Baseline Outcome Testing, Intervention Block 1 (weeks 1-4), Post-4 Week Blinded Outcome Testing, Intervention Block 2 (weeks 5-8), Post-8 Week Blinded Outcome Testing, Intervention Block 3 (weeks 9-12), and Post-12 Week Blinded Outcome Testing (Figure 2). To represent the cost per completed participant (n=15), the total cost for each chronological group was divided by 15.

## RESULTS

A total of 18 participants were enrolled at our site, with 15 completing the study. During the study, three participants withdrew due to adverse events.^16^ These participants withdrew after completing Post-4 Week Blinded Outcome Testing and were included in cost analysis for all visits completed. As shown in Figure 1, most participants lived within a 50km radius to the laboratory site. Two participants resided in rural, or frontier counties, which are indicated by light gray shading.

**Figure 1.**
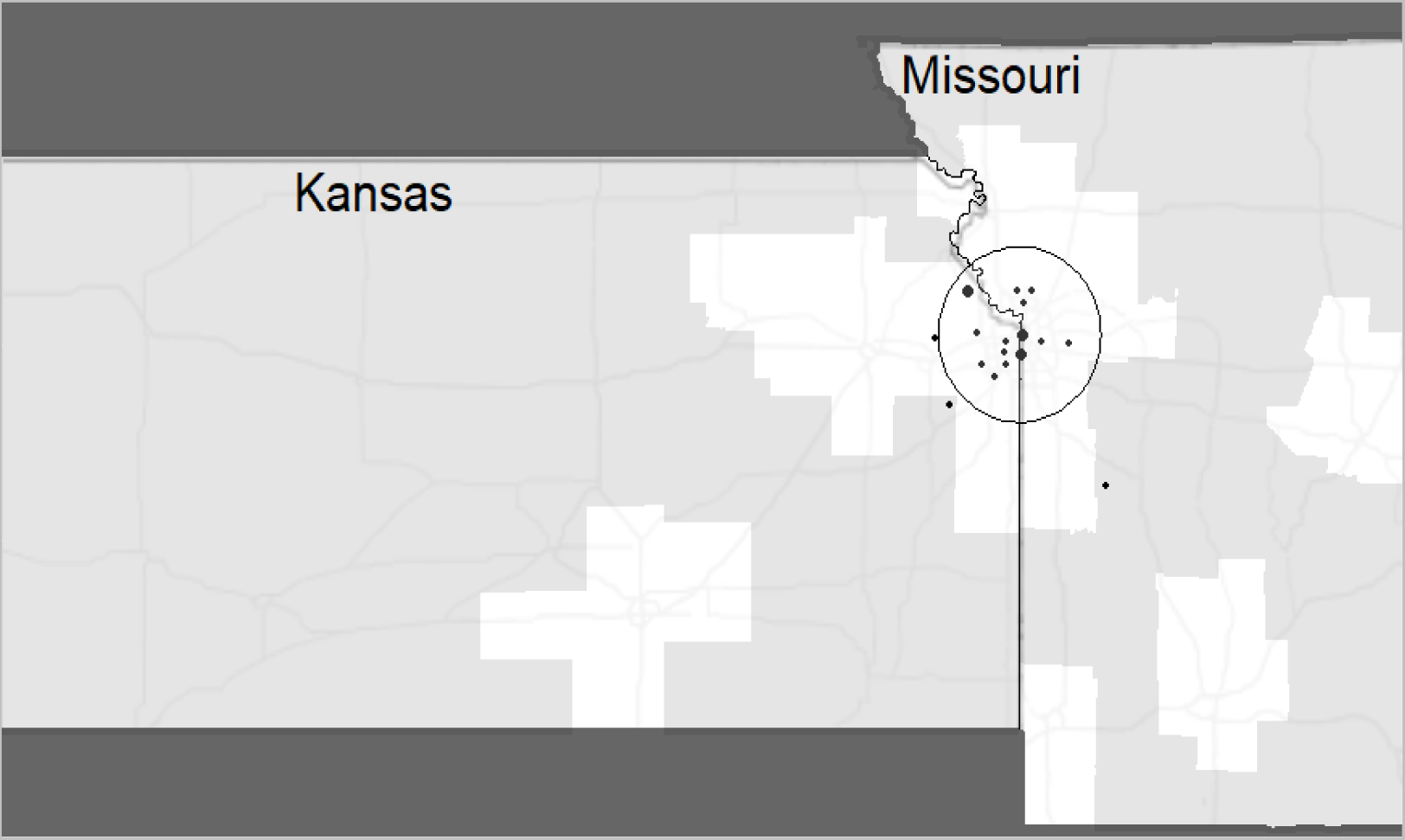
Distribution of participants enrolled in the HIT Stroke trial. The circle depicts a 50km radius to the laboratory with predominantly urban and suburban areas. Points are in arbitrary locations, equidistant to the laboratory to protect participant confidentiality. Point size indicates the number of participants recruited from the given area.

### TOTAL COST

The estimated total direct cost of the HIT Stroke Trial at the University of Kansas Medical Center was $539,768, resulting in a trial cost of $19,600 over the grant budget. This amount was offset using internal funds to support transportation costs that exceeded our site budget and to support a medical translator at each study visit and outcome testing. Figure 2 shows the percentages associated with the distribution of costs across oversight, recruitment, retention, and outcome assessment categories.

**Figure 2.**
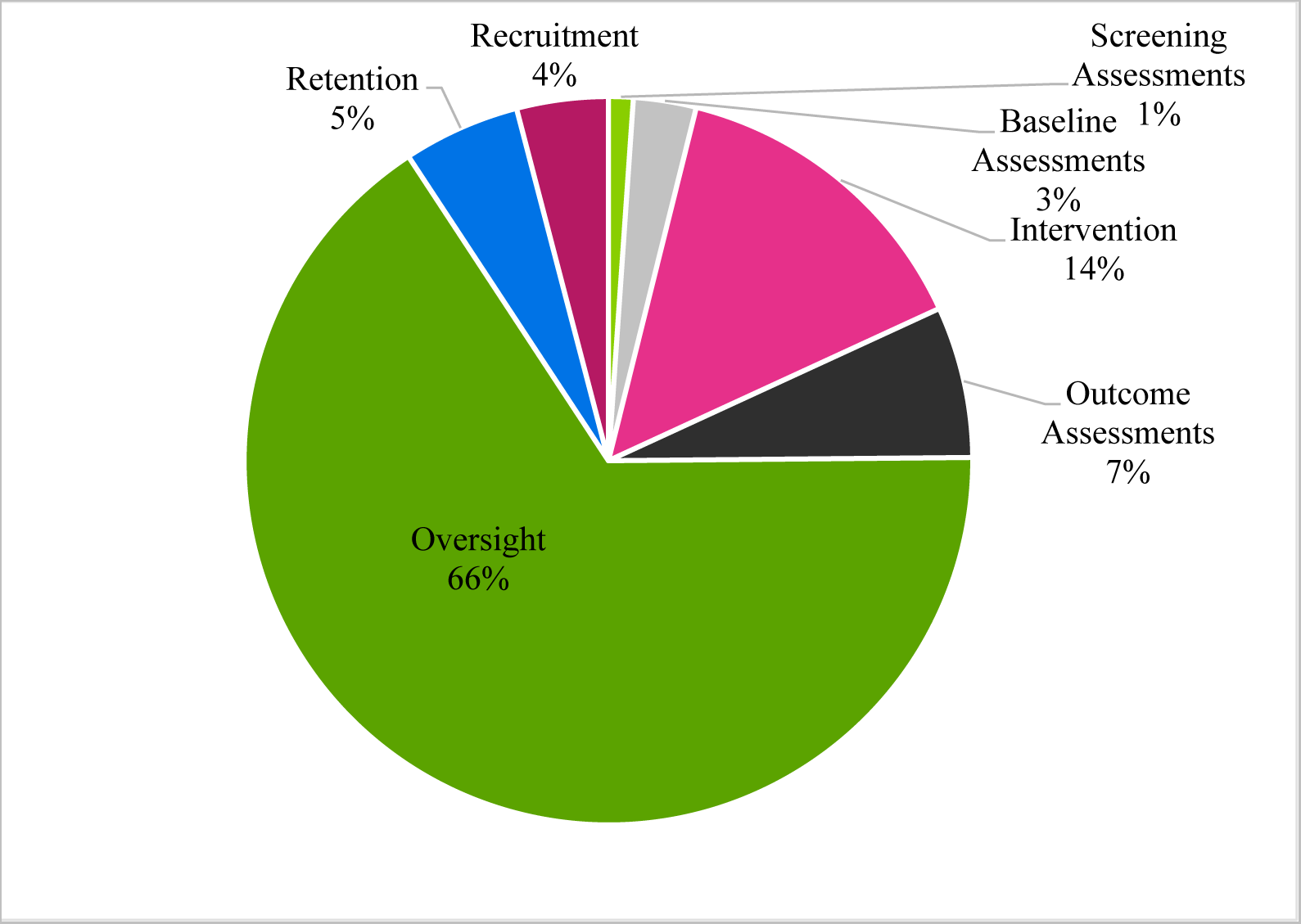
Percentages Associated with Trial Cost Distribution Across Categories

### COST PER PARTICIPANT

The overall cost for one enrolled participant to complete the study was estimated to be $35,984.54. This cost includes screening, consenting, baseline and outcome testing, and intervention sessions. Oversight and retention costs have been integrated into the following categories, capturing the commitment of both the principal investigator and study coordinator. Additionally, these categories encompass costs specifically allocated to enhance research accessibility. Figure 3 outlines the distribution of cost per participant.

**Figure 3.**
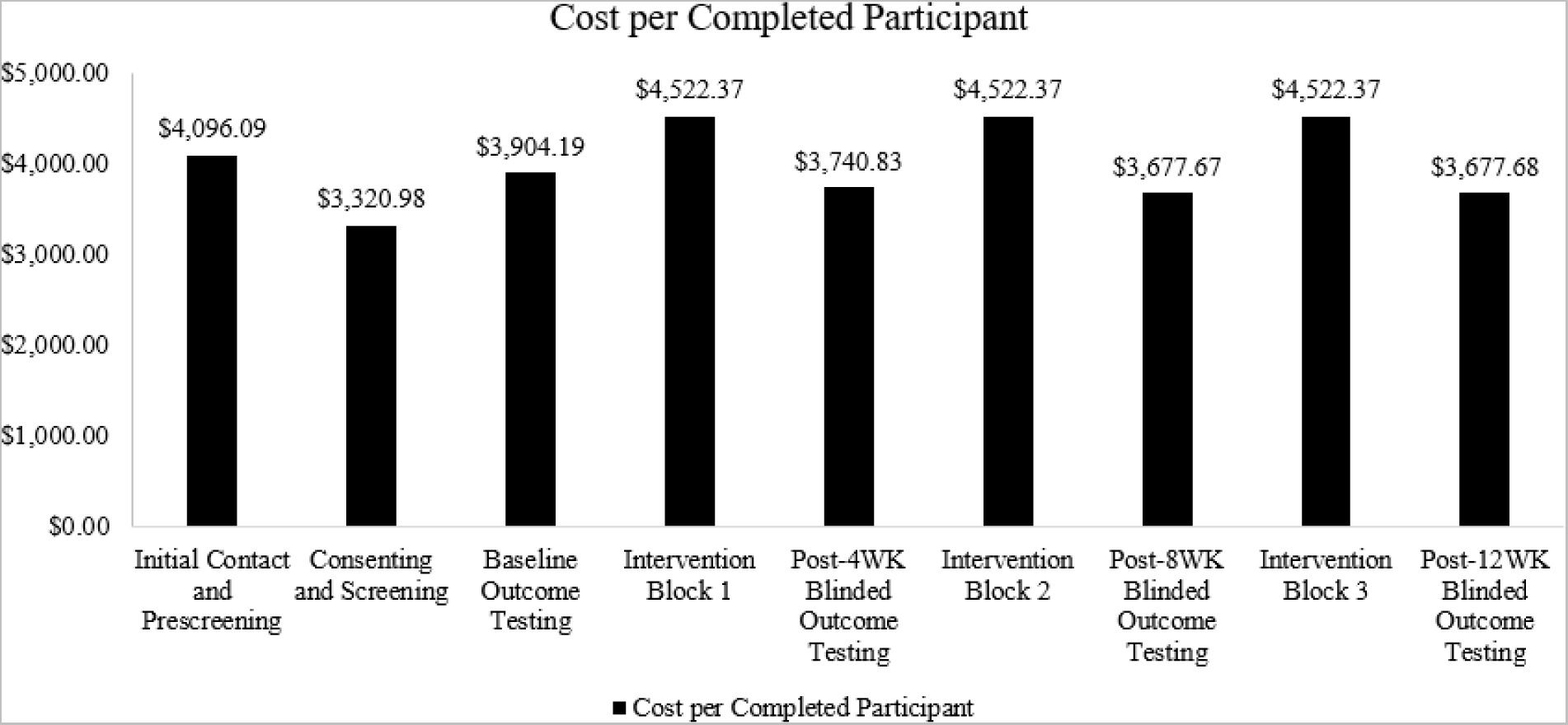
Cost for One Enrolled Participant to Complete Each Trial Phase

### RECRUITMENT

Recruitment costs totaled $21,923, including the cost of newspaper advertisements, media interviews, time spent introducing the trial in clinic and support groups, and staff time spent conducting phone screenings. As highlighted in Figure 3, cost is frontloaded per participant. This initial increased cost for one participant to complete the HIT Stroke trial occurs at Initial Contact and Prescreening and reflects the number of potential participants that must be contacted to enroll and randomize one participant. To determine the increased cost factor, defined as financial effort required to randomize one participant, we divided the number of participants randomized by total number of individuals phone screened to provide a randomization ratio.^13^ We then took the inverse of this ratio and established an increased cost factor of 5.88, which indicates that to randomize one participant, approximately six potentials had to be screened.^13^

### SCREENING and BASELINE ASSESSMENTS

Initial in-person screening assessments had an estimated cost of $5,905 which included physical therapist time and student assistants in the doctor of physical therapy program. Baseline assessment costs totaled $15,028. These assessments were completed at the University of Kansas Clinical and Translational Science Unit (CTSU) with a designated physical therapist, costing approximately $11,352. The cost for student assistants was approximately $3,675.

### INTERVENTION

The cost to conduct the HIT Stroke Trial intervention at the University of Kansas Medical Center was approximately $76,952. Training visits were completed by designated physical therapists, whose effort totaled at $49,756 and the student assistants resulted in a cost of approximately $3,675. Equipment needed for the intervention, including a treadmill, harness system, heart rate monitors, and iPods, had an approximate total cost of $23,521.

### OUTCOME ASSESSMENTS

Outcome assessments had an estimated cost of $36,288 and were completed at our University of Kansas CTSU by assessors blinded to group assignment. Outcome assessment costs included: 1) the bundled cost for the graded exercise tests with gas analysis and personnel. A certified exercise physiologist performed the graded exercise test and medical monitor read the electrocardiogram, 2) the physical therapist ensured participant safety with the treadmill and harness system throughout the graded exercise test, 3) the therapist performed all walking assessments; and 4) the study coordinator performed questionnaires.

### RETENTION

Figure 4 details the cost distribution related to participant retention. Retention costs totaled $28,009. Our site spent 76.1% ($21,334) on ride-share transportation to minimize the barriers of study participation. At study completion, all participants required transportation to a single study visit, highlighting the importance of providing transportation to minimize missed study visits or outcome testing. The primary reasons for transportation were: 1) family member unavailable, 2) car broke down, or 3) family member ill and unable to drive participant. Approximately, 25% of participants required transportation for all visits due to no transportation access including public transportation. We calculated ~$1,725 was spent on medical translation and interpreter services for one non-English speaking individual. For compensation, participants were provided $75 after each outcome testing visit, totaling $4,950. For study completion, participants were provided a completion certificate, for a total of $0.20.

**Figure 4.**
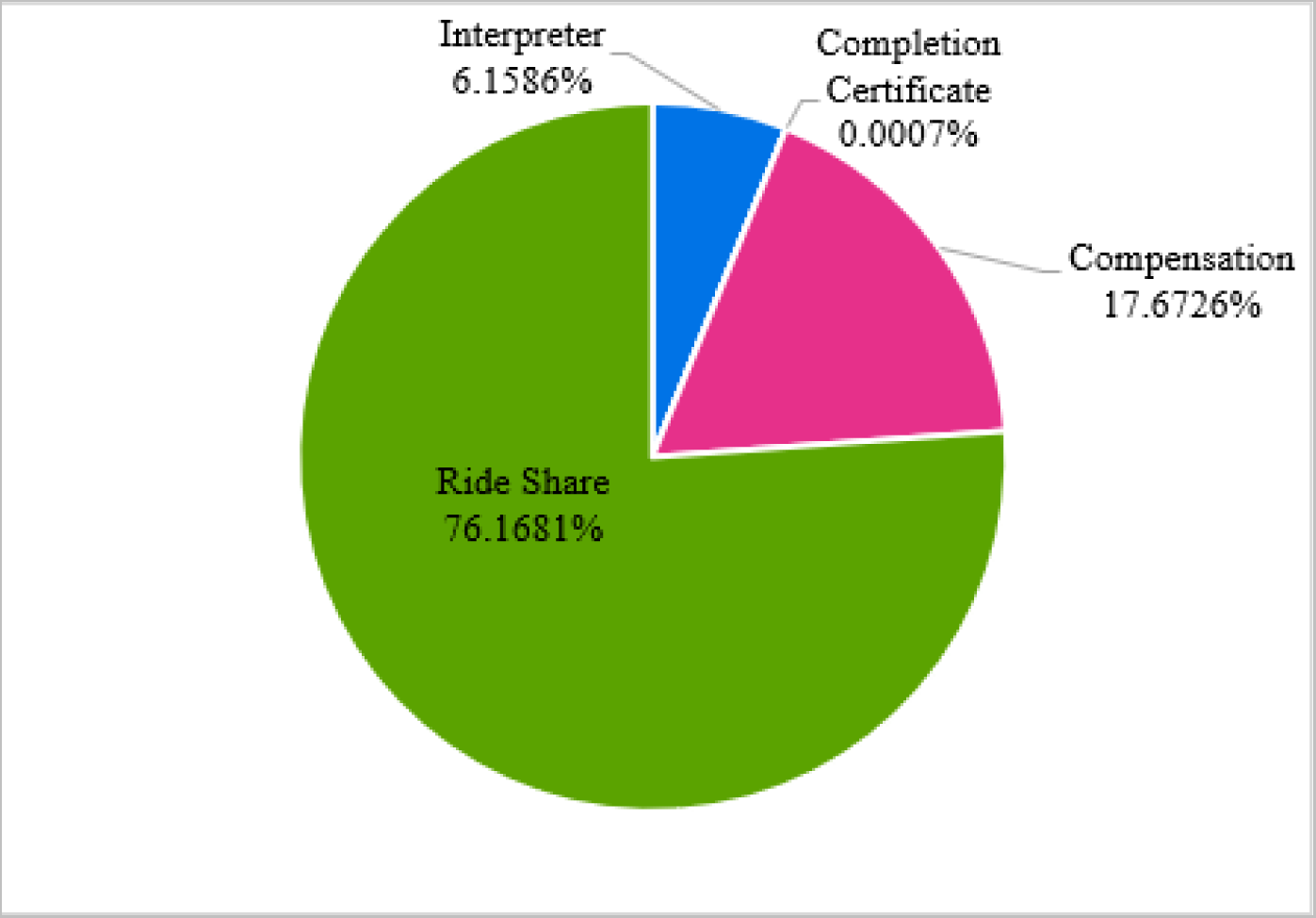
Cost of Participant Retention

### OVERSIGHT

Oversight costs for the trial duration totaled $355,661 and reflects principal investigator and study coordinator total compensation. Principal investigator effort was dedicated to all trial related activities including but not limited to budget, training study team members, personnel oversight, data monitoring, study meetings, creating annual reports for our site, and data safety and monitoring board preparation and meetings, totaling $215,273. Study coordinator salary was dedicated to managing regulatory documents including institutional review board, protocol adherence, assisting with consenting, adverse event documentation, screening and outcome assessments, and participant scheduling for a total of $140,388.

## DISCUSSION

This cost analysis aimed to elucidate the financial intricacies involved in recruitment, enrollment, outcome testing, and intervention for a clinical exercise trial focusing on stroke recovery. Understanding these costs is crucial for effective resource planning, budgeting, and fund procurement as stroke rehabilitation and recovery trials “are not simply acute stroke studies that are initiated at late time points”.^2^ Rather, the challenges associated with recruitment, retention, and intervention delivery are unique to stroke recovery and the information provided here should address potential costs and provide valuable information related to stroke recovery trials.

### INCREASED COST FACTOR AND RECRUITMENT

The data showed an average cost of enrollment and trial completion per participant was $35,984 with approximately 1/5 of cost dedicated to recruitment and screening for initial enrollment. The cost per participant is lower than an acute endovascular intervention where the cost of the intervention group was reported at $126,494 versus the control at $143,331.^26^ However, the costs reported in the acute endovascular study included resources used such as therapy during the 2-year follow up. For the present study, the increased cost factor indicated our site would conduct phone screens on at least 6 potential participants for one to be randomized. The increased cost factor reported here is similar to that observed in a recent 12-month exercise intervention trial in older adults, which reported an increased cost factor of 5.95 for phone screening and randomization for 494 participants.^13^ It is important to note, however, that factors such as study inclusion and exclusion criteria, as well as individual interest in a given study topic,^27^ may influence the increased cost factor. For example, a study with more strict inclusion and exclusion criteria may require increased participant screening for randomization, and these factors should be considered in study design. Further, successful recruitment methods may be implemented to minimize increased cost factor. We have previously published our approach for optimizing recruitment in stroke recovery trials, using a “service first” approach.^1^ Establishing supportive relationships with physicians and providing training to the recruitment team on effective participant communication before study start-up may optimize recruitment.^1^ Further, sites may consider using participant databases for a streamlined recruitment approach. Since completion of the HIT Stroke trial, our site has created a Stroke Recovery Registry in which we enroll participants interested in stroke recovery research. In our registry, we record demographics, medical history, and study interests to pair individuals who have experienced a stroke with potential research opportunities that meet their interests and needs based on inclusion and exclusion criteria to identify more quickly those who may be eligible during the phone screening phase.

### BASELINE AND OUTCOME ASSESSMENTS

Another driving factor for our site related costs in the HIT Stroke trial was baseline and outcome testing, including the personnel time dedicated to these measures. Testing included graded exercise tests, conducted by trained personnel with physical therapist oversight, physical function testing conducted by a physical therapist, and questionnaires conducted by the study coordinator. Graded exercise tests with gas analysis and a bodyweight support harness were conducted every 4 weeks for 12 weeks. The HIT Stroke study design focused on identifying both the intensity needed to change walking outcomes and the duration (4, 8, 12 weeks). Therefore, these additional outcome assessments added increased but necessary costs to the trial to answer the research questions. An additional consideration for baseline and outcome testing is assessor qualifications. The HIT Stroke trial employed licensed physical therapists for physical function assessments and added safety for the graded exercise testing. While utilizing physical therapists to conduct the study increases trial applicability for translation to clinical practice, necessitating an advanced degree and licensure in an assessor may increase assessment costs. Further, this requirement may decrease the generalizability of study results to settings without physical therapy availability.

### INTERVENTION

For exercise sessions, a treadmill and bodyweight support harness, heart rate and lactate monitors, iPods for real time monitoring of heart rate, and step watches for session step count were required. Personnel costs included licensed physical therapist to deliver the intervention and we employed doctor of physical therapy students to assist with the intervention. We acknowledge that our cost factor is likely influenced by the salaries associated with the need for licensed physical therapists. However, to ensure safe delivery of the intervention, we believe these costs are justified. The cost factor may not be identical to all locations across the United States and is likely dependent on regional cost of living and may differ between sites. For example, the cost of living index in the Kansas-Missouri region is 87.4-88.3, whereas it is 92.2 in Ohio, and 100.8 in Delaware.^28^ Further, the annual mean wage of physical therapists is higher in Ohio and Delaware than the Kansas-Missouri region.^29^ As such, staff salaries may differ between sites and needs to be considered when planning multi-site trials.

### PARTICIPANT RETENTION

#### Transportation

Our retention costs include expenses required to decrease barriers to research engagement. Approximately $21,334 were dedicated ride share services. Research suggests that transportation is a key barrier to research participation and results from decreased access to vehicles or public transportation, inadequate infrastructure, and travel distance and costs.^2,27,30,31^ Our site experiences challenges for participant transportation due to the poor availability of public transportation in the Kansas City Metro area. According to the 2021 American Community Survey^21^, 0.8% of individuals in the Kansas City Metro area use public transportation, and 86.3% use a car, truck, or van to commute to work. Conversely, at our partner sites in Newark, DE, and Cincinnati, OH, 3% to 6.2% of individuals use public transportation, and 69% to 77.5% use a car, truck, or van to commute to work, respectively. Additionally, due to geographics of the Kansas City Metro area, only 1.1% of individuals walk to work, whereas 11.6% and 5.5% in Newark and Cincinnati walk to work, respectively. Figure 1 shows the distribution of participants for the HIT Stroke trial in our catchment area and the willingness of people living with stroke to engage and participate in an exercise trial designed to improve walking. In advance of the clinical trial, we budgeted for participant transportation. However, our expenses related to transportation costs exceeded the budgeted amount in the grant. Internal funds and other sources of funding were used to support participants and reduce barriers to promote inclusive science.

In addition to transportation support, our team placed special emphasis on parking and navigating from parking locations to clinic, which have been cited as commonly neglected barriers to research engagement in stroke.^2^ Our site offered free parking to all participants in the parking site nearest our laboratory, which is less than 500 feet from the building entrance. We provided a detailed parking map and instructions prior to study visits. Additionally, if needed our staff would meet participants at their vehicles and provide wheelchair assistance to reduce fatigue prior to the study visit or assessment visits.

#### Translation services

To further decrease barriers to engagement at our site, we used other funding sources to cover costs of $1,725 to a medical translator and language interpreter to decrease barriers to clinical trial participation. Currently, over 7,000 languages exist worldwide, and the English predominance in research not only decreases research accessibility, but also the generalizability of research findings for individuals who do not speak English.^32^ Research suggests that these language barriers perpetuate healthcare inequities and may negatively impact healthcare policies and delivery.^32^ By providing transportation through ride-share programs and translator services, we minimize recruitment and retention barriers for all, create a trust-worthy environment within our academic medical center setting, and assist with place-based disparities for those in rural or urban settings without access to transportation.

#### Compensation

Participants were compensated for their time in the HIT Stroke trial. Although concerns have been raised regarding the potential for participant bias in studies which offer compensation,^33^ research suggests that compensation helps to promote participant retention^34^ and offset the financial burden placed on participants for study engagement.^35^ Participants often incur expenses as a result of study travel and time away from work for participation and providing study compensation can reduce socioeconomic disparities in research, where the burden of engagement is greater on those with lower income.^35^

#### Non-Financial Incentives for Participation

Lastly, we provided participants with a certificate of study completion. Research suggests that non-financial incentives which express appreciation for participation are commonly used in studies and help promote retention.^34^ Due to financial limitations in clinical trials, consideration of non-financial incentives may be optimal for helping to reduce participant drop out and loss to follow-up bias.^34^

These data provide novel findings related the cost associated with conducting a stroke recovery trial. While we tracked study related costs, we acknowledge that the cost per participant and increased cost factor are estimates, as it is impossible to determine how many individuals were reached through study advertisements or actual effort was spent on each category such as phone screens, emailing or calling participants and coordinating with the ride-share programs. Further, we report costs for a single site of the HIT Stroke trial. Costs may differ between sites due to factors such as staff salaries based on regional cost of living, costs for space usage, transportation needs, and translation services. We did not account for costs that may have been associated with operations during the COVID-19 pandemic including personal protective equipment, additional staffing, screening participants for COVID-19, time lost due to staff or participants who reported COVID-19 exposure and institution approved cleaning solutions to protect against COVID-19. Additional costs that were not accounted for at our site additional staff for data checking and time spent on coordinating delivery of equipment and supplies and the set-up of equipment. Finally, we acknowledge these costs are associated with trial execution in the United States. Therefore, some costs may not be relevant to all locations across the world.

## Conclusion

This analysis outlines the costs associated required to conduct a single blind randomized exercise trial focused on stroke recovery. Here, we provide a model for increasing participation and retention to enroll participants effectively and inclusively for stroke recovery trials aligned with our “service first” approach. We believe that this cost analysis will provide investigators and funding agencies with a valuable information regarding the costs required to conduct a successful and accessible exercise-based clinical trial in stroke.

## Data Availability

Data is available upon request.

## Acknowledgments

The authors would like to thank Sasha Moores and Saniya Waghmare for their assistance with gathering cost information. The content is solely the responsibility of the authors and does not necessarily represent the official views of the NIH.

## Non-standard Abbreviations and Acronyms

HIT: High-Intensity Interval Training
MAT: Moderate Aerobic Training
PHQ-9: Patient Health Questionnaire 9
EQ-5D: EuroQol-5D
ABC: Activities-specific Balance Confidence Scale
PROMIS: Patient Reported Outcomes Measurement Information System Fatigue Scale,^24^
GXT: Graded Exercise Test
CTSU: Clinical and Translational Science Unit

---

**Table 1.** Increased Cost Factor Associated with Recruitment and Screening.

| Assessment | Participants<br>Assessed | Participants<br>Randomized | Randomization<br>Ratio | Increased Cost<br>Factor |
| --- | --- | --- | --- | --- |
| Phone Screening | 106 | 18 | 0.17 | 5.88 |

